# α1 adrenoreceptor antagonism mitigates extracellular mitochondrial DNA accumulation in lung fibrosis models and in patients with IPF

**DOI:** 10.1101/2022.04.06.22273471

**Authors:** Genta Ishikawa, Xueyan Peng, John McGovern, Sam Woo, Carrighan Perry, Angela Liu, Sheeline Yu, Alexander Ghincea, Huanxing Sun, Changwan Ryu, Erica L. Herzog

**Author notes:** Equal contribution (Changwan Ryu, Erica L. Herzog). To whom correspondence should be addressed <u>Correspondence:</u> Erica L. Herzog, M.D., Ph.D., 300 Cedar Street TAC 441S, New Haven CT 06520-8057.

## Abstract

Idiopathic Pulmonary Fibrosis is increasingly associated with adrenergic innervation and endogenous innate immune ligands such as mitochondrial DNA (mtDNA). Interestingly, a connection between these entities has not been explored. Here we report that noradrenaline (NA) derived from the lung’s adrenergic nerve supply drives the accumulation of αSMA-expressing fibroblasts via a mechanism involving α1 adrenoreceptors and mtDNA. Using the bleomycin model of lung fibrosis we compared the effect of lung specific adrenergic denervation achieved via the inhalational administration of the sympathetic neurotoxin 6-hydroxydopamine to surgically mediated adrenal ablation and found that NA derived from local but not adrenal sources drives lung fibrosis. Bleomycin induced the appearance of a αSMA+ fibroblast population co-expressing the adrenoreceptor alpha-1D (ADRA1D). Therapeutic delivery of the α1 adrenoreceptor antagonist terazosin reversed these changes and suppressed the accumulation of extracellular mtDNA. TGFβ1-stimulated normal human lung fibroblasts treated with TGFβ1 and Noradrenaline expressed ADRA1D and developed reduced αSMA expression and extracellular mtDNA concentrations when treated with terazosin. IPF patients prescribed α1 adrenoreceptor antagonists for non-pulmonary indications showed improved survival and reduced concentrations of plasma mtDNA. These findings link nerve-derived NA and α1 adrenoreceptor antagonism with mtDNA accumulation and lung fibrogenesis in mouse models, cultured cells, and humans with IPF. Further study of this neuro-innate connection may yield new avenues for investigation in the clinical and basic science realms.

## INTRODUCTION

Idiopathic pulmonary fibrosis (IPF) is a progressive and incurable disease defined by the presence of usual interstitial pneumonia (UIP) in the absence of an identifiable cause (1). The median survival is at best 2-4 years. Most patients die from respiratory failure due to disease progression and disease incidence and prevalence are increasing (1–3). While currently available treatment options slow the rate of lung function decline, treatment responses are highly variable, incredibly costly, and associated with at times intolerable side effects (4, 5). Because lung transplantation is the only curative treatment option, IPF remains a chronic, fatal disease for which there is an urgent need for reasonably priced therapies that safely and effectively modify the clinical course.

Pulmonary fibrosis is proposed to originate from interactions between alveolar epithelial injury and dysregulated innate immunity that through poorly understood mechanisms engender accumulation of alpha smooth muscle actin (αSMA)-expressing myofibroblasts and excessive extracellular matrix (ECM) (6–8). One factor that has been little studied in this regard is the role of nerve-derived signals. While the lung’s autonomic nerve supply has been effectively targeted for treatment of airway disease (9), far less is known about nerve-lung interactions in parenchymal disorders. It is thus relevant that interventions targeting parasympathetic (10) and sympathetic innervation (11) have shown efficacy in mitigating fibrotic endpoints in animal models of IPF, with the latter being at least partially due to suppression of adrenergic processes driven by Noradrenaline. Noradrenaline (NA), an adrenergic catecholamine that exerts its effect on target cells via G protein coupled receptors of the alpha and beta families (12), is increasingly implicated in the inflammatory remodeling of bone marrow (13), adipose tissue (14), gastrointestinal tract (15), tumor microenvironment (16), and in some settings, lung (4, 17–19). Our work (11) has shown that fibrotic lungs are enriched for ectopically patterned adrenergic nerves, that these nerves drive fibrosis by releasing NA, and that NA-driven fibrosis can be treated with α1 adrenoreceptor antagonists. This axis is active in IPF where lung tissues are enriched for markers of adrenergic innervation and NA, and patients treated with α1 adrenoreceptor antagonists experience improved clinical outcomes. These clinical observations are exciting yet limited in not having determined how lung NA accumulates and the mechanism(s) through which α1 adrenoreceptor antagonism mitigates fibrotic endpoints in animal models and improves outcomes in patients with IPF. Better understanding of the source of lung NA and the mechanism of its associated lung remodeling will be required for the safe and effective targeting of these processes in patients with IPF.

Innate immunity has emerged an intervenable amplifier of fibrotic pathologies in the lung and other organs (8). Innate immune activation is stimulated by pattern recognition receptors (PRRs) that recognize epitopes broadly conserved across microbial pathogens, known as pathogen-associated molecular patterns (PAMPs); and endogenous ligands, known as danger-associated molecular patterns (DAMPs), that are released by injured or activated cells and tissues (20). These responses occur both in professional immune cells and in non-immune populations such as fibroblasts. In fact, our group (21) and others (22) have demonstrated the relevance of mitochondrial-derived DAMPs such as mitochondrial DNA (mtDNA) (23) in a series of reports indicating that this endogenous ligand for cytosolic DNA sensors drives the autocrine activation of αSMA+ myofibroblasts (21). Interestingly, despite NA’s increasingly appreciated connection to mitochondrial DNA integrity (24), a direct relationship to extracellular mtDNA accumulation remains obscure and benefit for α1 adrenoreceptor inhibition – which improves mitochondrial DNA stability in other conditions (25) – has not been assessed. Improved understanding of this relationship will impart better understanding of the mechanism through which these agents mitigate inflammatory fibrosis and illuminate a new form of neuro-innate crosstalk that could be leveraged for clinical benefit.

In this study, we combine animal modeling, cell culture studies, and primary human biospecimens to show that NA derived from local adrenergic nerves induces lung fibrosis via a mechanism involving αSMA+ cells that express α1-adrenoreceptors and mtDNA, and that this axis is at least partially active in patients with IPF.

## METHODS

### Study approval

All animal experiments were performed according to the principles approved by the Yale University IACUC with handling adhering to the Guide for the Care and Use of Laboratory Animals. All human studies were performed with approval from institutional human investigation committee at Yale University, and written informed consent was received from participants at the time of enrollment into our repository.

### Animals

Wild type male and female C57BL/6J mice obtained from The Jackson Laboratory were used between age 7 and 9 weeks. Bleomycin administration (2.0U/kg), bronchoalveolar lavage (BAL), and lung harvest were performed according to our previous description (26). Mice that had undergone adrenalectomy or sham surgery were purchased from The Jackson Laboratory and provided 0.9% saline in their drinking water to prevent cardiovascular collapse due to mineralocorticoid deficiency as is commonly done in studies of this type (27). Unless stated otherwise, groups were evenly divided between males and females.

### Chemical denervation of lung adrenergic nerves via inhalational route

Animals were anesthetized by placing them in a chamber with paper towels soaked with 40 percent isoflurane solution diluted with 1,2-propanediol, followed by orotracheal administration of 500 μg of 6-hydroxydopamine (6-OHDA, dissolved in 50 μl of sterile PBS containing 0.1% ascorbic acid; Sigma-Aldrich) or vehicle (0.1% ascorbic acid) per mouse daily for 3 days (28).

### Terazosin treatment

Mice were randomly divided into two groups five days after the administration of bleomycin. One group was intraperitoneally injected with 1 mg/kg terazosin from day 5 to day 13; another group was synchronously treated with PBS delivered via the same route to serve as vehicle controls. Mice were euthanized on day 14.

### Sacrifice and lung harvest

After animals were terminally anesthetized, BAL was performed followed by median sternotomy and right heart perfusion. The lungs were then processed for the molecular and biochemical assays described previously (29).

### Bronchoalveolar lavage cell quantification

BAL cell counts were determined as previously described (26).

### Quantification of mouse BAL cytokine concentrations

BAL fluid was subject to multiplex quantification of IL-1β, IL-6, IP-10, MCP-1, and TNFα, using the U-PLEX Biomarker Group 1 (ms) Assays (Meso Scale Discovery, Rockville, MD, Catalog Number: K15069L-1) per the manufacturer’s protocol. Briefly, BAL samples were diluted in a 1:5 ratio and plated in duplicate. Cytokine concentrations were quantified and presented in pg/mL using the Meso Scale Discovery QuickPlex SQ 120 Model 1300 with Discovery Workbench 4.0 software.

### Collagen quantification

Lungs were snap frozen in liquid nitrogen and stored at −80°C until they underwent quantification of soluble collagen using the Sircol Collagen Assay kit (Biocolor Ltd., CLS 1111) per manufacture’s protocols (29).

### Histologic analysis

Whole left lungs were harvested from experimental mice for histological analysis. Formalin fixed, paraffin embedded sections were stained with Masson’s trichrome to visualize collagen deposition (30). The Modified Ashcroft Score (MAS) was performed with a minor modification of previously published method (31). This quantitative scoring system assigns a numerical value between 0 and 8 that correspond to the extent of fibrosis in a histologic sample.

### Immunofluorescence and fluorescence microscopy

Immunofluorescence detection of α1 adrenoreceptors on mouse lung sections was performed using rabbit monoclonal ADRA1A (1:100, Abcam, ab137123), rabbit monoclonal ADRA1B (1:1000, Abcam, ab169523), rabbit polyclonal ADRA1D (1:100, Abcam, ab84402), and mouse anti–αSMA (1:250, Abcam, ab7817) antibodies followed by secondary detection and DAPI counterstaining. Negative controls included slides that were processed and detected in the absence of primary antibody.

### Immunofluorescence for imaging lung adrenergic nerves

Vibratome sectioning and immunofluorescence staining were carried out in which the rate-limiting enzyme in the biosynthesis of catecholamines, tyrosine hydroxylase (TH), was detected as a adrenergic neuron marker. Lung preparation for vibratome sectioning was performed as previously described (32). The 150-μm-thick lung sections were blocked overnight with 5% normal goat serum (NGS) in 0.5% Triton X-100/1× PBS (PBS-T) at 4°C on a shaker. Sections were then incubated with rabbit anti-TH (1:100, Abcam, ab112) and mouse anti–αSMA (1:250, Abcam, ab7817) antibodies for 3 days at 4°C. After washing with PBS-T, lung sections were continuously incubated overnight with the corresponding Alexa Fluor–488 goat anti–rabbit IgG (1:500, Invitrogen, A11008) and Alexa Fluor 555–goat anti–mouse IgG (1:500, Invitrogen, A21422) for 24 hours at 4°C. Finally, lung sections were washed 4 times with PBS-T and mounted with the VECTASHIELD antifade mounting medium containing 1.5 μg/mL DAPI (Vector Laboratories, H-1200). Images were taken using an SP8 confocal laser microscope with the software of Leica Application Suite X (Leica Microsystems Inc.). Negative controls included slides that were processed and detected in the absence of primary antibody.

### Determination of noradrenaline and adrenaline concentrations

Concentration of NA and adrenaline (A) was determined in serum, BAL, lung and liver tissues using commercially available NA and A high-sensitivity ELISA kits (DLD Diagnostika, GMBH, NOU39-K01 and ADU39-K01) that we have used for this application in other publications (11).

### DNA isolation

DNA was extracted from 200 μl BAL and cell culture supernatants or 100 μl plasma using the QiaAMP DNA Mini-Kit (Qiagen, Germantown, MD) per their protocol.

### Human mtDNA quantification for cell culture supernatant and plasma samples

The presence of mtDNA in supernatant and plasma samples was assayed by real time quantitative polymerase chain reaction (qRT-PCR) for the human MT-ATP6 gene, using commercially available primers for MT-ATP6 (ThermoFisher Scientific, Catalog number: 4331182) and probes (SsoAdvanced Universal Probes Supermix, Bio-Rad Laboratories, Hercules, CA). The ViiA 7 Real-Time PCR System (ThermoFisher Scientific) was employed with the following cycling conditions: incubation at 95° for 10 minutes followed by 50 amplification cycles of 95° for 10 seconds, 60° for 30 seconds, and 72° for 1 second, and finally cooling at 40° for 30 seconds (33). The number of MT-ATP6 copies per microliter (copies/µl) was determined based on a standard curve developed from serial dilutions of a commercially available DNA plasmid (OriGene, Rockville, MD) with complementary DNA sequences for human MT-ATP6.

### Mouse Mitochondrial DNA quantification for BAL

The presence of mtDNA in BAL samples from mice was assayed by qRT-PCR for the mouse COX1 gene, and probes (SsoAdvanced Universal SYBR Green Supermix, Bio-Rad Laboratories). The ViiA 7 Real-Time PCR System (Roche, Indianapolis, IN) system was employed with the following cycling conditions: incubation at 95° for 10 minutes followed by 40 amplification cycles of 95° for 10 seconds, 55° for 3 seconds, and 72° for 10 seconds and melting curve stage of 95° for 1 second, 65° for 15 seconds, and 95° for 15 seconds. The number of COX1 copies per microliter (copies/µl) was determined based on a standard curve developed from serial dilutions of a commercially available DNA plasmid (OriGene) with complementary DNA sequences for mouse COX1. As a negative control we utilized primers for 16s rRNA to amplify any commensal bacteria that maybe present in the mouse BAL.

The primers for mouse samples were:

COX1-F: 5′-GCCCCAGATATAGCATTCCC-3′;

COX1-R: 5′-GTTCATCCTGTTCCTGCTCC-3′;

*27*-F: 5′-AGAGTTTGATCCTGGCTCAG-3 ′;

*1492*-R: 5′-GGTTACCTTGTTACGACTT-3 ′;

### RNA isolation and qRT-PCR

Total cellular RNA was isolated from normal human lung fibroblasts (NHLF) using trizol based methods as previously described (29). RNA was reverse transcribed and evaluated for expression of αSMA (*ACTA2*) and *GAPDH* as appropriate. Relative expression was quantified using the 2delta Ct method as we have previously reported (29). Data were reported as fold change relative to baseline.

Primers for NHLF samples were as follows:

*ACTA2*-F: 5′-GTGTTGCCCCTGAAGAGCAT-3′;

*ACTA2*: 5′-GCTGGGACATTGAAAGTCTCA-3′;

*GAPDH*-F: 5′-TGGAGAAGGCTGGGGCTCATTT-3′;

*GAPDH*-R: 5′-TGGTGCAGGAGGCATTGCTGAT-3′;

### Culture and TGF-β1/noradrenaline stimulation of normal human lung fibroblasts

NHLFs obtained from Lonza (Allendale, NJ) were stimulated with and without 50 µmol/l of NA in the presence or the absence of 10 μmol/L of terazosin and 5 ng/ml of recombinant human TGFβ1 as previously described (34).

### FACS

FACS of cultured NHLFs was performed on an LSRII flow cytometer. The supernatants were dumped and Accutase (EMD Millipore Corp) was used to detach cells. After centrifuging, the pellets were resuspended in FACS buffer (1× PBS containing 1% FBS, 0.01% NaN3, and 1 mM EDTA) at a density of 1 × 10^6^ cells/mL. Next, cells were incubated with rabbit polyclonal ADRA1D (1:100, Abcam, ab84402) diluted in FACS buffer containing 1% NGS for 1 hour at 4°C. Where relevant, secondary detection of the unconjugated primary antibody was then performed (AF700, goat anti-rabbit, Invitrogen, A21038). Finally, cells were washed with FACS buffer and filtered through a 100-μm cell strainer. Acquisition was performed on an LSRII flow cytometer using FACSDiva acquisition software. FlowJo software (FlowJo, LLC) was used for data analysis. The positive and negative gates were set based on relevant isotype control.

### MTT assay

NHLF viability was assayed with the 3-(4,5-dimethylthiazol-2-yl)-2,5-diphenyltetrazolium bromide (MTT) assay (ThermoFisher Scientific) per manufacturer’s protocol. Briefly, NHLFs were detached with 0.05% trypsin/EDTA, counted, and approximately 10,000 cells were seeded onto 96-well cell culture plates. Cells were labeled with 12 nM MTT, lysed with 100 ul of DMSO (99.9%), and analyzed at an absorbance of 570 nm with the Vmax Kinetic Microplate Reader (Molecular Devices, Sunnyvale, CA) with SoftMax Pro 5.4 software.

### Statistical Analyses

All data are presented as means ± SEM or median ± IQR unless stated otherwise. Normally distributed data were compared using 1- or 2-tailed student’s *t* test or ANOVA. Non-normally distributed data in two groups were compared using the nonparametric Mann-Whitney test. Categorical data were compared with one-way analysis of variance and Fisher’s exact test. Receiver operating characteristic curve (ROC) analysis of the Yale-ILD cohort was used to determine the mtDNA cutoff. Kaplan-Meier analysis was performed to determine survival associations. Multivariate Cox regression hazard ratios (HRs) were used to model associations. GraphPad Prism 9.0 (GraphPad Software, La Jolla, CA) was used for all these analyses. A p-value <0.05 was significant.

## RESULTS

### Local adrenergic nerves are required for bleomycin induced lung fibrosis

Nerve-derived Noradrenaline (NA) is increasingly implicated as a cross-organ regulator of inflammatory pathology (35) and our own work in this area found that inhibition of NA accumulation via chemical and genetic methods suppressed bleomycin induced pulmonary fibrosis (11). However, because these methods ablated adrenergic nerves in a ubiquitous fashion, a direct contribution of lung innervation could not be assessed. Similarly, because the methods were by necessity constitutive, they could only be tested in a preventative approach. Hence, to improve the rigor of prior studies and more firmly link fibrosis to nerve-derived NA, we adapted a recently published model of lung denervation (28). For these studies, mice randomized to receive 3 successive doses of 500 μg 6-OHDA administered via the orotracheal route displayed a lung-specific reduction in TH+ nerve content, and tissue NA, that was significantly reduced from vehicle-treated animals and not evident in extrapulmonary organs such as the liver (Figure S1).

Having confirmed this method as a powerful tool to ablate adrenergic nerves in a lung specific fashion, we then questioned whether this intervention protects lungs from experimentally induced fibrosis. Here, bleomycin challenged mice were randomized to receive inhaled 6-OHDA on a therapeutic schedule from day 5 through 7 post-bleomycin and then followed for an additional 7 days at which point they were sacrificed and relevant endpoints assessed (Figure 1, A). Evaluation of adrenergic nerve content assessed by tyrosine hydroxylase (TH+) immunostaining revealed near absence of these structures in denervated lungs (Figure 1, B-C). Further evidence of successful denervation was provided when 6-OHDA treated lungs displayed a specific reduction in NA (which enters the lung from both nerves and the circulation), but not Adrenaline (A) (which is predominantly produced in the adrenal glands) (Figure 1, D and E). These changes were accompanied by a significant reduction in fibrotic endpoints such as collagen content (Figure 1, F), Modified Ashcroft Scores (MAS) (Figure 1, G), and trichrome staining (Figure 1, H-I). Importantly, inflammatory mediators that may be induced by 6-OHDA were unaltered, as BAL white blood cell (WBC) count and inflammatory cytokine concentrations (IL-1β, IL-6, IP-10, MCP-1, and TNF-α) were similar between 6-OHDA and vehicle treated groups (Figure S2). These findings support the concept that intrapulmonary adrenergic nerves are required for maximal fibrosis in this model.

**Figure 1:**
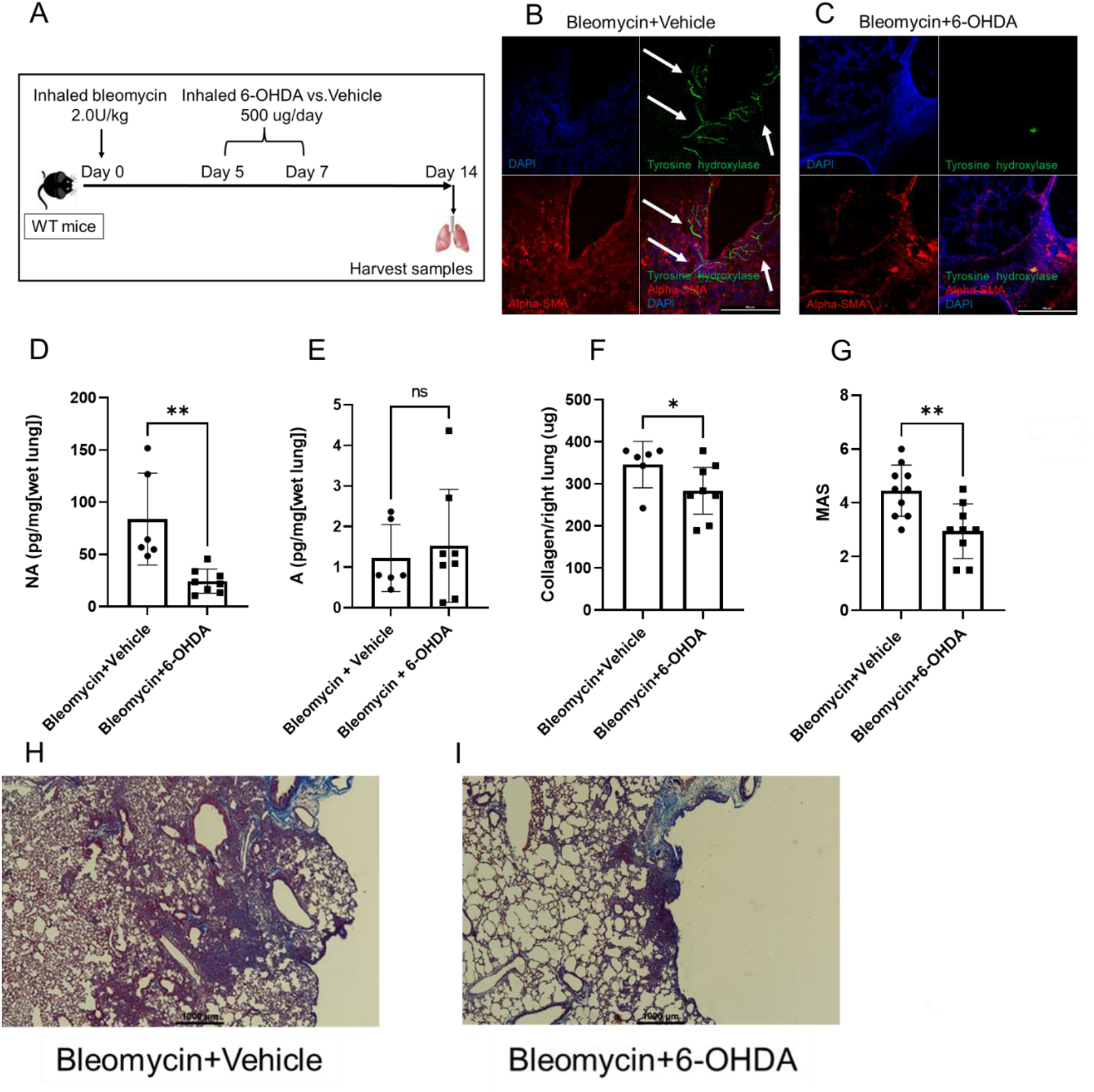
Inhaled 6-OHDA ameliorates experimentally induced lung fibrosis. (**A**) WT mice challenged with inhaled bleomycin (2.0U/kg) on Day 0 were treated with either L-ascorbic acid (vehicle) or inhaled 6-OHDA from days 5 through 7 and sacrificed at the 14-day time point. (**B** and **C**) Immunofluorescence detection of αSMA (red), TH (green), and DAPI (blue) in bleomycin-challenged lungs treated with vehicle (**B**), and bleomycin-challenged lungs treated with 6-OHDA (**C**). In the bleomycin + vehicle image in **B**, white arrows indicate TH+ linear structures in parenchymal regions. In the bleomycin + 6-OHDA image in **C**, TH+ linear structures are undetectable in this region. (**D** and **E**) Comparison of noradrenaline (NA) and adrenaline (A) concentrations (pg/mg[wet lung tissue]) between in mice challenged with vehicle (left) and inhaled 6-OHDA (right) revealed that relative to vehicle-challenged groups, the NA concentration was substantially decreased at the 14-day time point in mice challenged with inhaled 6-OHDA (**D**, *P* = 0.0030), but the A concentration was unchanged (**E**, *P* = 0.6466). (**F**-**I**) Relative to mice with vehicle (left), mice with inhaled 6-OHDA (right) showed reduced right lung collagen (**F**, *P* = 0.0409) and, in a separate experiment, both MAS (**G**, *P* = 0.0040) and trichrome staining (**H**, “Bleomycin + Vehicle” and **I**, “Bleomycin + 6-OHDA”). \**P* < 0.05, \*\**P* < 0.01. Data are shown as mean ± SEM. A: adrenaline. α-SMA: α-smooth muscle actin. BAL: bronchoalveolar lavage. MAS: Modified Ashcroft Score. NA: noradrenaline. 6-OHDA: 6-hydroxydopamine. TH: tyrosine hydroxylase. WT: wild-type. In **H** and **I** original images are 10X original magnification. Scale bar = 100 microns.

### Adrenal glands are dispensable for bleomycin induced lung fibrosis

The data above compellingly demonstrate a requirement for NA derived from local nerves in the development of experimentally induced lung fibrosis. They do not, however, rule out the contribution of NA or A derived from additional sources such as adrenal glands. To address this gap in knowledge we employed a well-accepted model of bilateral adrenal ablation achieved by surgical means. This approach, which has been widespread for decades, involves the surgical resection of adrenal glands (or sham surgery) followed by the administration of saline-supplemented drinking water to prevent cardiovascular collapse from adrenal failure (27). Twenty-one days post-surgery mice received a single inhaled dose of 2.0 U/kg of pharmacological grade bleomycin (Figure 2, A) and were followed for an additional 14 days at which point they were sacrificed and assessed for fibrotic endpoints. We first confirmed the near total absence of circulating A (Figure 2, B), indicating that the intervention was successful in ablating adrenal-derived catecholamines. Interestingly, the reduction in NA approached but did not meet statistical significance (Figure 2, C) likely reflecting its production in other locations. Consistent with our prediction, the absence of adrenal glands failed to impact lung inflammation measured by BAL cell counts, collagen accumulation measured by the Sircol Assay, and histologic evidence of fibrosis reflected by MAS and trichrome staining (Figure 2, D-H). These observations indicate that adrenal glands are dispensable for bleomycin induced lung fibrosis and suggest that targeting locally derived NA may be more beneficial in this model.

**Figure 2:**
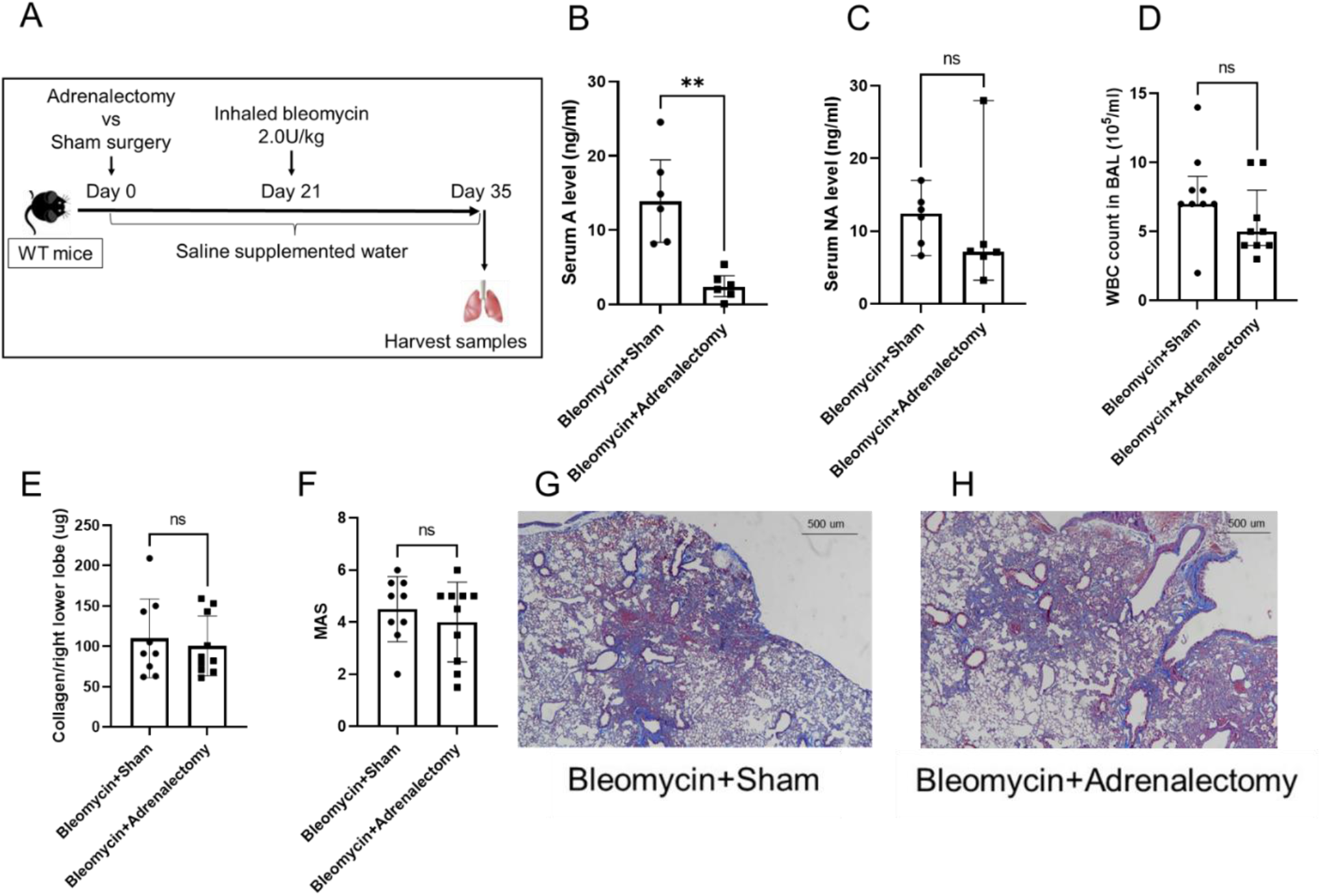
Adrenal glands are dispensable for bleomycin induced lung fibrosis. (**A**) WT mice that underwent bilateral adrenalectomy or sham surgery were challenged with inhaled bleomycin (2.0U/kg) after 3 weeks of postsurgical recovery period during which mice were given saline supplemented drinking water. They were then sacrificed at the 14-day time point after bleomycin administration. (**B** and **C**) Comparison of the serum adrenaline (A) and noradrenaline (NA) level in adrenalectomized mice (right) challenged with bleomycin, relative to mice with sham surgery (left), revealed serum A level was significantly decreased (**B**, *P* = 0.0011), but NA (**C**, *P* = 0.1797) level was unchanged. (**D**) Mice with adrenalectomy showed unchanged WBC count in BAL (right), relative to mice with sham surgery (left) (*P* = 0.1022). (**E**-**H**) Relative to mice with sham surgery (left), mice with adrenalectomy (right) showed unchanged right lower lobe collagen (**E**, *P* = 0.6372), MAS (**F**, *P* = 0.4489) and trichrome staining (**G**, “Bleomycin + Sham” and **H**, “Bleomycin + Adrenalectomy”). \*\**P* < 0.01. Data are shown as mean ± SEM or median ± IQR as appropriate. A: adrenaline. BAL: bronchoalveolar lavage. MAS: Modified Ashcroft Score. NA: noradrenaline. WT: wild-type. Figure **G** and **H** are 20X original magnification. Scale bar = 500 microns.

### ADRA1D expressing αSMA+ cells accumulate in fibrotic lung parenchyma

Next, we evaluated the cell(s) and receptor(s) through which nerve-derived NA drives fibrosis in this model. In light of NA’s relatively high affinity for α1 adrenoreceptors (12) and our published work in which α1 adrenoreceptor blockade mitigates fibrosis (11), we assessed expression of these proteins in the lungs of mice that did and did not receive bleomycin treatment. Here, detection of Adrenoceptor Alpha 1A (ADRA1A) and Adrenoceptor Alpha 1B (ADRA1B) were both low at baseline and after bleomycin appeared to localize to airway epithelial regions (Figure 3, A-B) with little to no staining in parenchymal regions. In contrast, staining with Adrenoceptor Alpha 1D (ADRA1D), which was also low at baseline, demonstrated robust signal in fibrotic regions at the 14-day timepoint following bleomycin (Figure 3, C). To ascertain if ADRA1D expression might identify fibrotic lung fibroblasts, we performed colocalization with αSMA, a marker that is commonly used to identify myofibroblasts in this setting. Here, unchallenged lungs showed the expected position of αSMA+ cells around airways and large vessels with minimal signal in parenchymal regions and no co-expression of ADRA1D (Figure 3, D and data not shown). Conversely, bleomycin exposed lungs showed the expected increase in parenchymal αSMA+ cells which displayed significant co-localization of ADRA1D (Figure 3, E-G). Specificity of this observation was shown by an absence of signal for either ADRA1A or ADRA1B in αSMA+ cells (Figure S3). These findings suggest that nerve-derived NA drives experimentally induced lung fibrosis through a mechanism involving ADRA1D-expressing αSMA+ cells.

**Figure 3:**
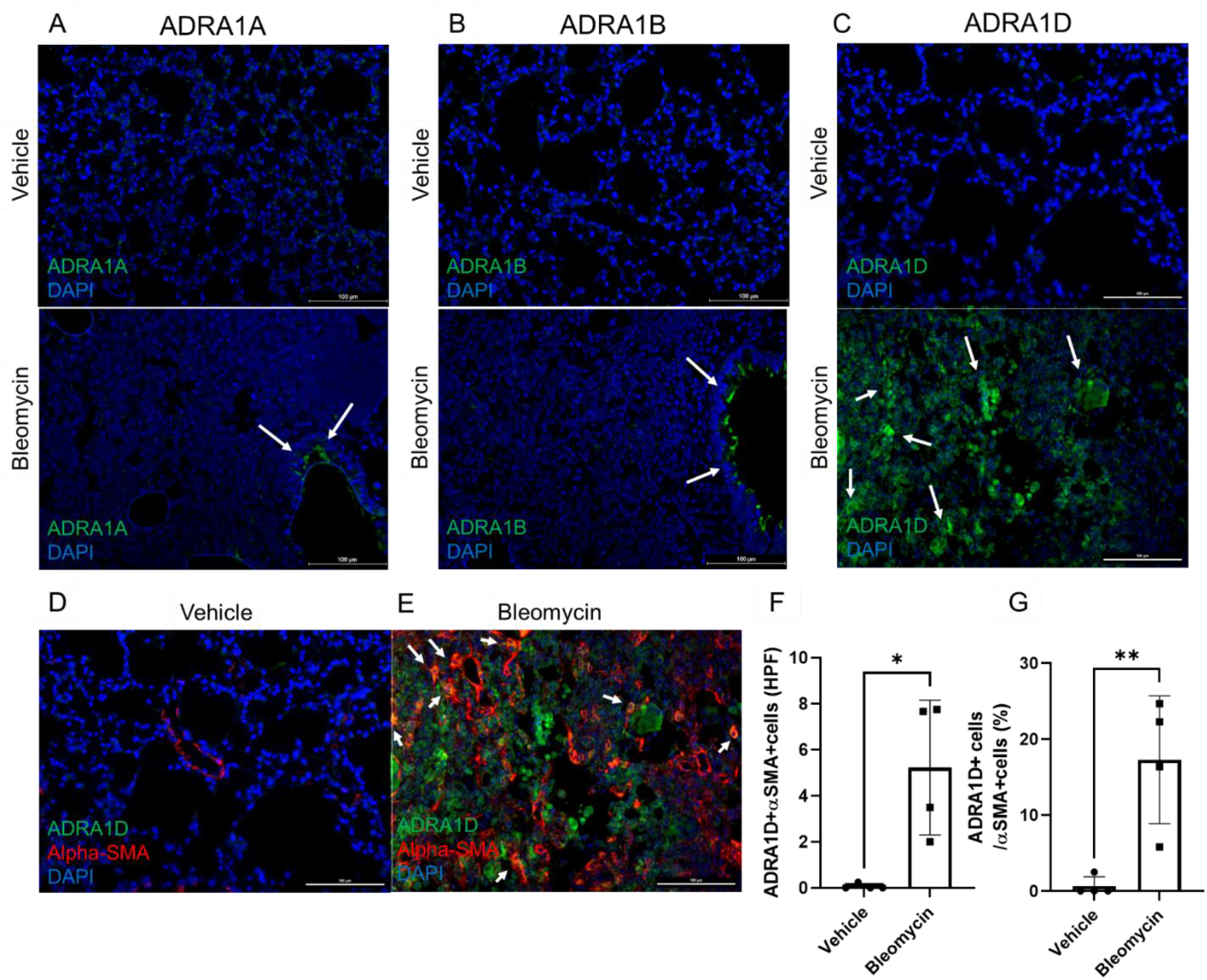
Geospatial localization of cells expressing ADRA1D and αSMA in bleomycin challenged lungs: WT mice were challenged by inhalational bleomycin (2.0U/kg) or vehicle on Day 0 and lung samples were harvested on Day 14. (**A** and **B**) Immunofluorescence detection of ADRA1A or ADRA1B (green) and DAPI (blue) in the bleomycin challenged lungs revealed staining with ADRA1A (**A**) or ADRA1B (**B**) was low at baseline and after bleomycin appeared to localize to airway epithelial regions (white arrows). (**C**) Immunofluorescence detection of ADRA1D (green) and DAPI (blue) in the bleomycin challenged lungs revealed robust signal in fibrotic regions (white arrows) at the 14-day timepoint following bleomycin. (**D** and **E**) Immunofluorescence detection of ADRA1D (green), αSMA (red), and DAPI (blue) in bleomycin challenged lungs revealed that vehicle challenged lungs (**D**) showed no cells co-expressing αSMA and ADRA1D, while bleomycin exposed lungs (**E**) contained cells co-expressing αSMA and ADRA1D + in fibrotic (white arrows). ADRA1D+ cells that do not co-express αSMA were also seen. (**F** and **G**) Relative to control lung tissue (left), there were a significantly higher quantities (**F**, *P* = 0.0124) and frequencies (**G**, *P* = 0.0078) of αSMA+ ADRA1D+ cells in the bleomycin-challenged lung tissues (right). \**P* < 0.05, \*\**P* < 0.01. Data are shown as mean ± SEM. ADRA1: α1 adrenoreceptor. αSMA: α-smooth muscle actin. HPF: high-power field. WT: wild-type. Scale bar = 100 microns.

### α1 adrenoreceptor antagonism reduces accumulation of parenchymal αSMA+ cells

In our previous work we showed that therapeutic dosing of terazosin, an α1 adrenoreceptor antagonist with relative high affinity for ADRA1D, reduced bleomycin-induced lung collagen accumulation and remodeling (11), but the mechanism of this observation remains unknown. Because ADRA1D was detected on the parenchymal αSMA+ fibroblasts that arise as a relatively late event in fibrosis, we questioned whether terazosin’s therapeutic benefit involved this population. To this end, we replicated our prior study in which bleomycin treated mice were randomized to receive 1mg/kg of terazosin from days 5-14 at which point they were sacrificed and samples were assessed (Figure 4, A). Consistent with our prior studies, terazosin significantly reduced collagen accumulation as measured by the Sircol assay (Figure 4, B). Additionally, relative to mice treated with vehicle (Figure 4, C), mice treated with terazosin showed a marked and reduction in αSMA+ cells in the bleomycin induced fibrotic lung parenchyma (Figure 4, D). These data show that the antifibrotic mechanism of α1 adrenoreceptor antagonism extends to a reduction in αSMA+ cells.

**Figure 4:**
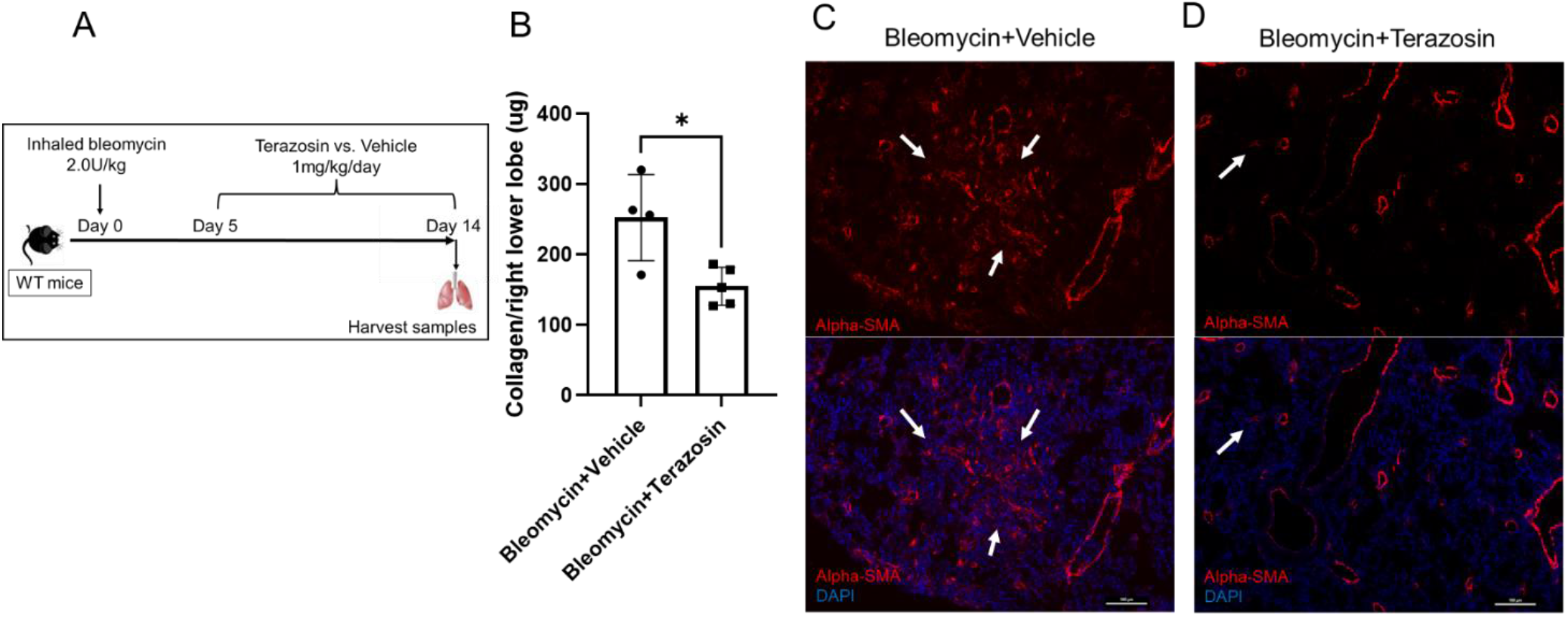
Terazosin reduces lung fibrosis in bleomycin induced lung fibrosis with decreased αSMA positive cells: **(A)** Bleomycin-challenged mice were randomized to receive therapeutic dosing of terazosin or vehicle control in a regimen of daily dosing initiated at day 5 and continued until the mice were euthanized at day 14. (**B**) Relative to bleomycin-challenged mice treated with vehicle (left), mice treated with terazosin (right) showed decreased lung collagen (*P* = 0.0144). (**C** and **D**) Immunofluorescence detection of αSMA (red) and DAPI (blue) in bleomycin-challenged lungs treated with vehicle (**C**), and bleomycin-challenged lungs treated with terazosin (**D**). In the vehicle lung (**C**), the parenchymal location of αSMA expressing cells (white arrows) in response to bleomycin challenge was observed. Relative to mice treated with vehicle, mice treated with terazosin showed reduced αSMA expression (white arrow) in this region (**D**). \**P* < 0.05. Data are shown as mean ± SEM. αSMA: α-smooth muscle actin. WT: wild-type. Scale bar = 100 microns.

### α1 adrenoreceptor antagonism reduces extracellular mtDNA accumulation

Numerous mechanisms have been implicated in the expansion of fibrotic lung fibroblasts expressing αSMA. One such process involves the activation of cytosolic DNA sensors by DAMPS, particularly DNA derived from mitochondria (mtDNA) that is released by stressed or injured cells into the extracellular milieu (21, 36). Given terazosin’s increasingly appreciated role in the stability of mitochondria and their genome (37), we assessed whether terazosin’s suppression of αSMA+ cell expansion was related to the accumulation of extracellular mtDNA, BAL samples from bleomycin challenged mice that did or did not receive therapeutic terazosin dosing were evaluated for mtDNA content using quantitative PCR for the mouse-specific mitochondrial gene COX1. (38). To ensure the rigor of these findings we first established the standard curve for these primers (Figure 5, A) and confirmed that they do not amplify commensal bacteria present in the mouse lung by using 16S rRNA, housekeeping bacterial genetic marker (39) (Figure 5, B). Next, as compared to bleomycin-challenged mice treated with vehicle, BAL specimens from animals treated with terazosin demonstrated substantially reduced mtDNA copy number (Figure 5, C). Taken together, these findings demonstrate that the antifibrotic effects of α1 adrenoreceptor antagonism with terazosin include the previously unrecognized suppression of mtDNA accumulation in the extracellular space.

**Figure 5:**
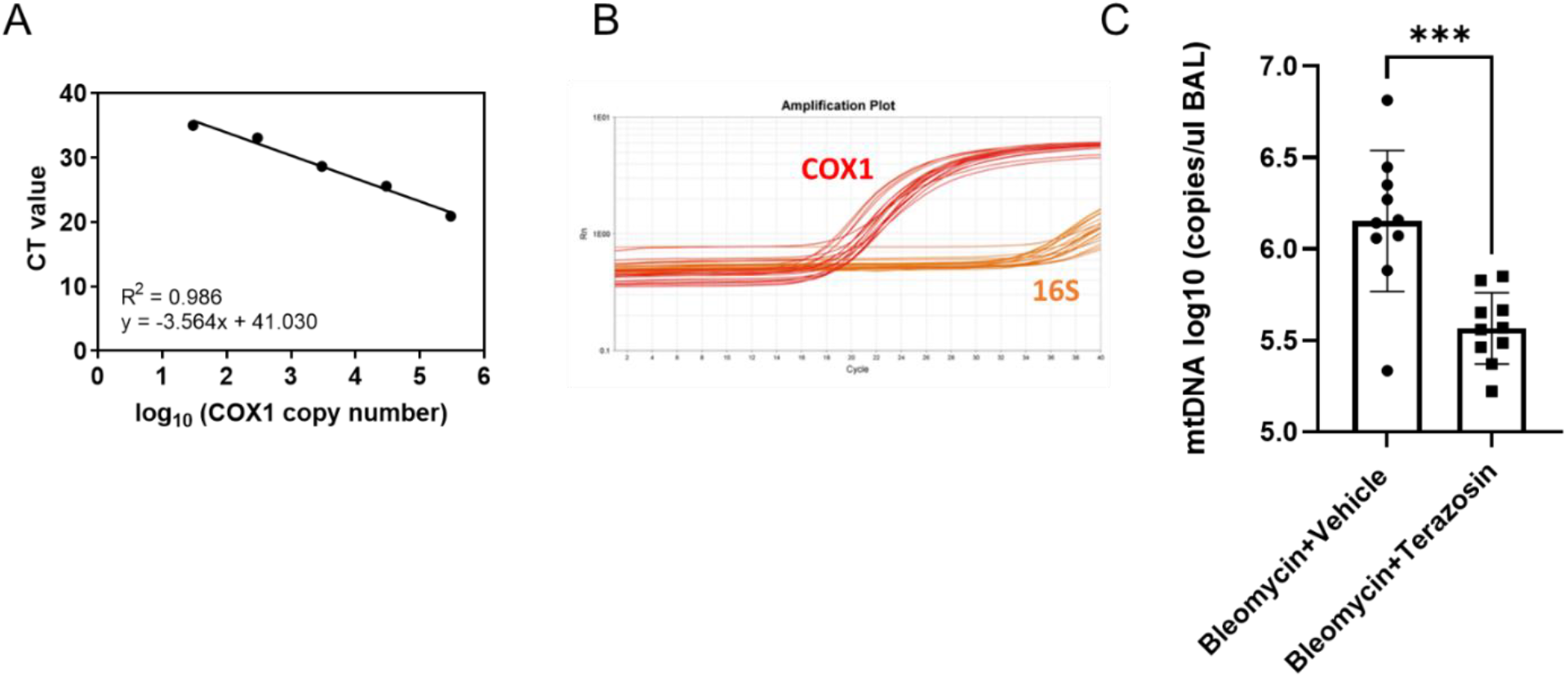
Terazosin reduces extracellular mtDNA accumulation in the bleomycin challenged murine lung. **(A)** The standard curve of COX1 copy number and CT value in qRT-PCR. (**B**) The amplification curve of COX1 and 16S rRNA. **(C)** Relative to BAL obtained from bleomycin-challenged mice treated with vehicle (left), a significant decrease in COX1 concentration was detected in BAL from animals treated with terazosin (right) (*P* = 0.0004). Data are presented graphically as log base 10 of the raw values (COX1 copies per microliter of BAL) with mean (±SEM). \*\*\**P* < 0.001. BAL: bronchoalveolar lavage. CT: cycle threshold. mtDNA: mitochondrial DNA. 16S: 16S rRNA.

### α1 adrenoreceptor antagonism exerts cell-autonomous suppression of αSMA expression and mtDNA release in TGFβ1 stimulated human lung fibroblasts

The *in vivo* findings of ADRA1D+ fibroblasts combined with terazosin-mediated reductions in αSMA expression and extracellular mtDNA accumulation led us to probe a relationship between these entities *in vitro*. First, we questioned whether terazosin’s benefit was mediated by a cell-autonomous, receptor-mediated process in fibroblasts. We first confirmed that NHLFs display surface expression of ADRA1D in the presence or absence of TGFβ1 stimulation (Figure 6, A). We then investigated whether this process was associated with fibroblast activation by assessing *ACTA2* expression on cell lysates and extracellular mtDNA release in culture supernatants using the well-validated method of quantifying the human mtDNA specific gene, MT-ATP6 (40, 41). NHLFs were grown to confluence and co-stimulated with 5 ng/mL TGFβ1 and 50 μmol/L of NA in the absence or presence of 10 µmol/L of terazosin, which significantly reduced the relative expression of *ACTA2* (Figure 6, B) and concentrations of extracellular MT-ATP6 (Figure 6, C) similar to our *in vivo* experiments. These findings were unlikely due to alterations in viability, as assessment of this endpoint via the MTT assay was similar across all conditions (Figure 6, D). Taken together, these data show that terazosin mitigates *ACTA2* expression and extracellular mtDNA accumulation in a cell-autonomous manner.

**Figure 6:**
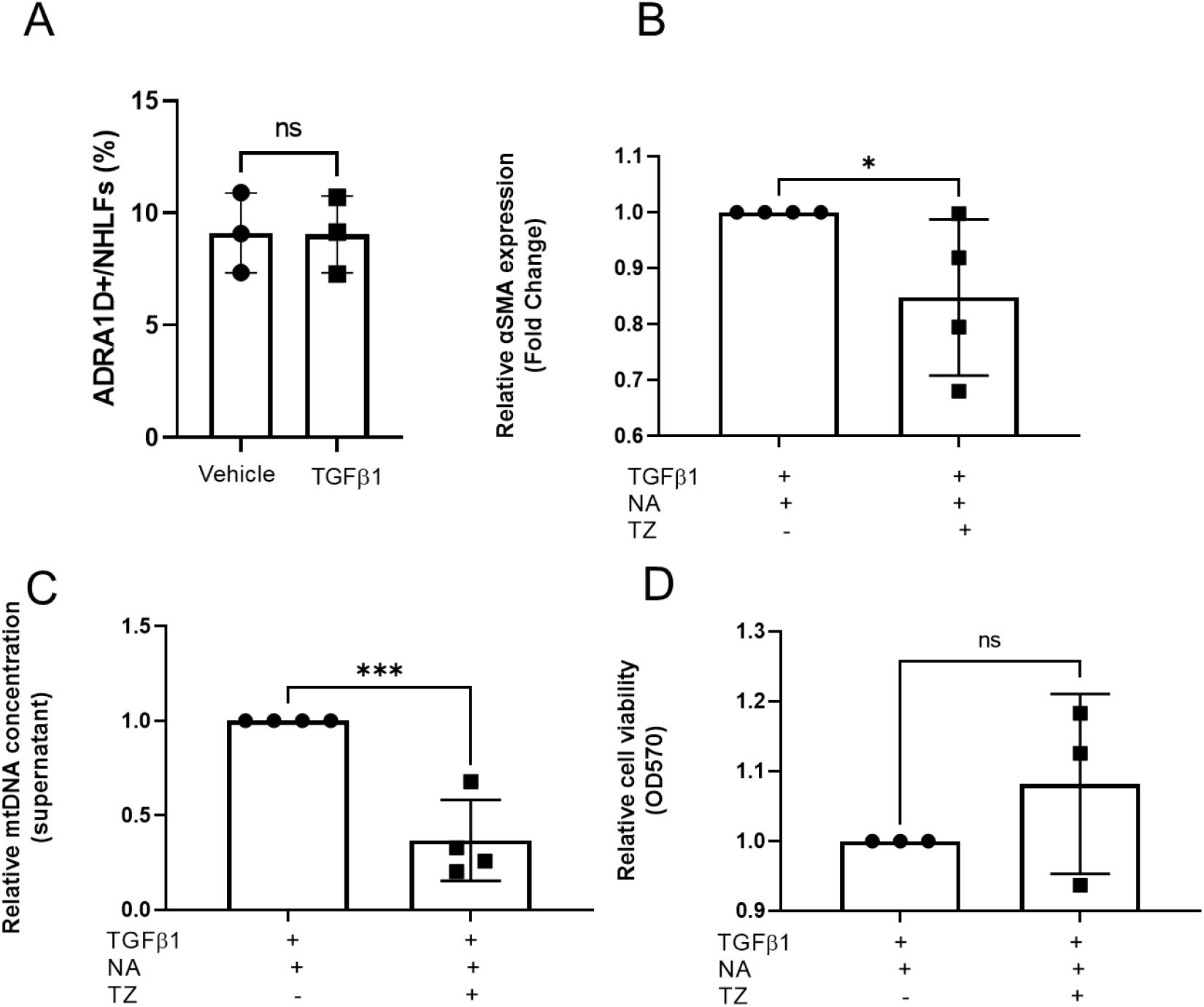
Terazosin reduces αSMA expression and extracellular mtDNA concentration in normal human lung fibroblasts. (**A**) Flow cytometric detection of ADRA1D+ fibroblasts in single-cell suspensions prepared from primary cell lines of normal human lung fibroblasts (NHLFs) challenged with vehicle (left) or human recombinant TGFβ1 (5 ng/mL for 48 hours, right) in vitro. Left axis shows percentages of fibroblasts expressing ADRA1D. Percentages of ADRA1D-expressing cells were independent of extrinsic human recombinant TGFβ1 challenge (*P* = 0.9631). (**B**) Treatment with terazosin (TZ) for NHLFs stimulated by 5 ng/mL of human recombinant TGFβ1 and 50 umol/L of noradrenaline (NA) for 24 hours significantly decreased the relative expression of αSMA (*P* = 0.0357). (**C**) Compared to cell culture supernatants obtained from NA and TGFβ1-stimulated NHLFs untreated with TZ (left), the relative concentration of MT-ATP6 was significantly decreased in cell culture supernatants obtained from NHLFs treated with TZ (right) (*P* = 0.0005). (**D**) Assessment of cell viability with the 3-(4,5-dimethylthiazol-2-yl)-2,5-diphenyltetrazolium bromide (MTT) assay revealed unchanged absorbance at a wavelength of 570 nm in NHLFs treated with TZ (*P* = 0.1663). \**P* < 0.05, \*\*\**P* < 0.001. Data are shown as mean ± SEM. ADRA1: α1 adrenoreceptor. αSMA: α-smooth muscle actin. mtDNA: mitochondrial DNA. MTT: the 3-(4,5-dimethylthiazol-2-yl)-2,5-diphenyltetrazolium bromide. NA: noradrenaline. NHLF: normal human lung fibroblast. TZ: terazosin.

### α1 adrenoreceptor antagonism decreases plasma mitochondrial DNA concentrations in IPF patients

α1 adrenoreceptor antagonism is emerging as a potential treatment for diseases characterized by inflammatory remodeling such as ARDS and COVID-19 (11, 42, 43) and we have previously shown that IPF patients taking these agents experienced preserved lung function and improved survival (11). In an attempt to link these clinical observations to the basic science studies shown above, we evaluated the relationship between plasma mtDNA concentrations – a reproducible biomarker of both fibroblast activation and mortality in the IPF disease state (21, 44, 45) – and α1 adrenoreceptor antagonism t in a longitudinal cohort of IPF patients followed at the Yale ILD Center of Excellence. First, we assessed mtDNA concentrations in plasma samples from IPF patients who were (n=12) or were not (n=32) taking α1 antagonists for a non-pulmonary indication (Table 1). In this small cohort of patients, Kaplan Meier analysis confirmed both the predictive value of plasma mtDNA copy numbers exceeding ATP6 copy number of 3.785 log10 for all-cause mortality in IPF (Figure 7, A and S4) along with the survival benefit of α1 adrenoreceptor antagonism in IPF (Figure 7, B). To assess whether these observations were linked, we compared plasma MT-ATP concentrations between these groups. Here, as expected, compared to patients not prescribed α1 adrenoreceptor antagonists, IPF patients prescribed this class of medications showed a significant reduction in plasma MT-ATP6 concentrations (Figure 7, C). These data show that the apparent survival benefit of α1 adrenoreceptor antagonism in IPF patients involves a reduction in circulating mtDNA concentrations.

**Table 1:**
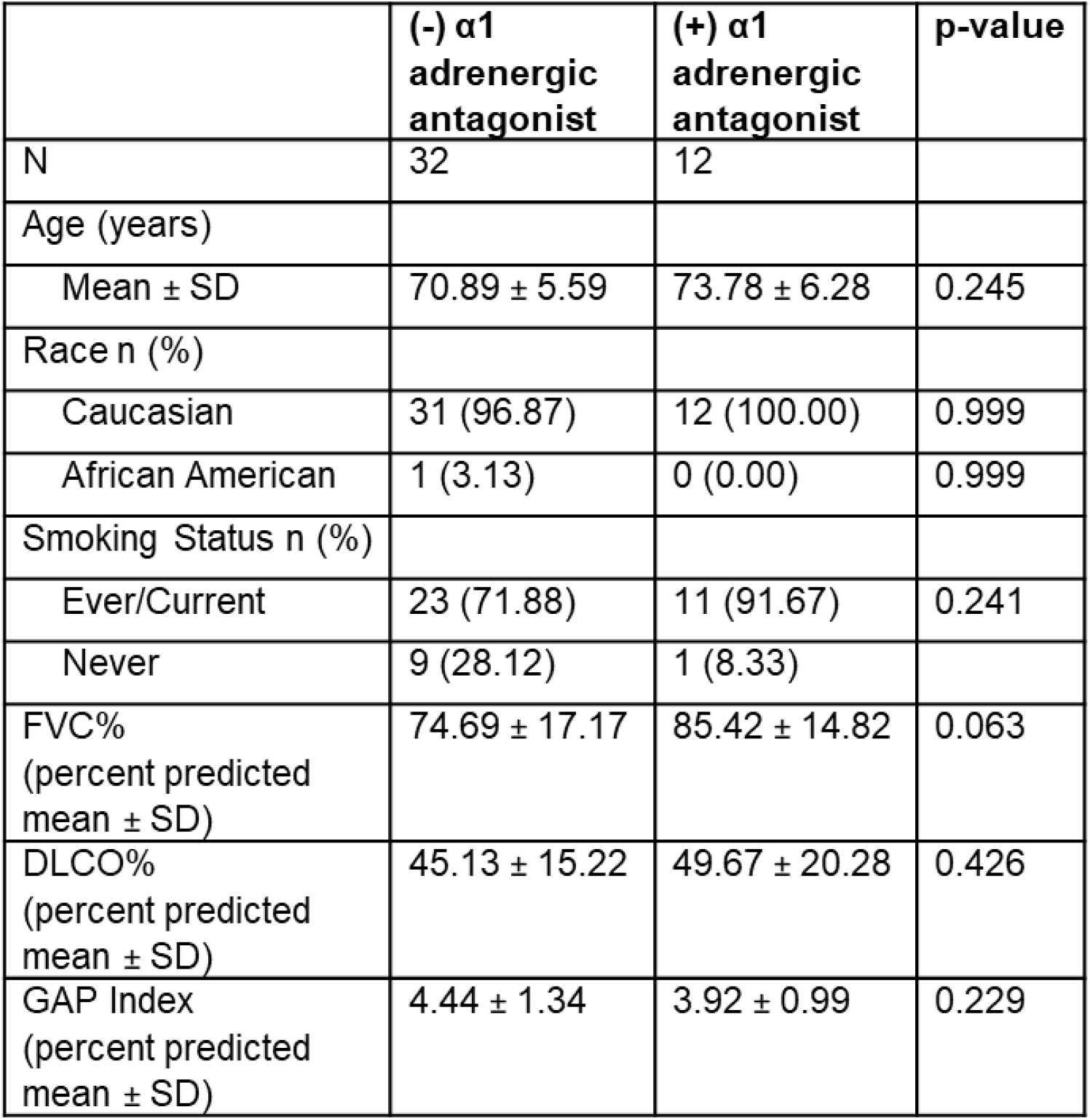
Clinical characteristics of the IPF population. Clinical characteristics between two groups were compared with *P*-values. Normally distributed data were compared using student’s *t* test or ANOVA. Non-normally distributed data were compared using the nonparametric Mann-Whitney test. Categorical data were compared with one-way analysis of variance and Fisher’s exact test. FVC%: percent predicted forced vital capacity. DLCO-%. Percent predicted diffusion capacity of carbon monoxide. GAP: Gender, Age, Physiology.

**Figure 7:**
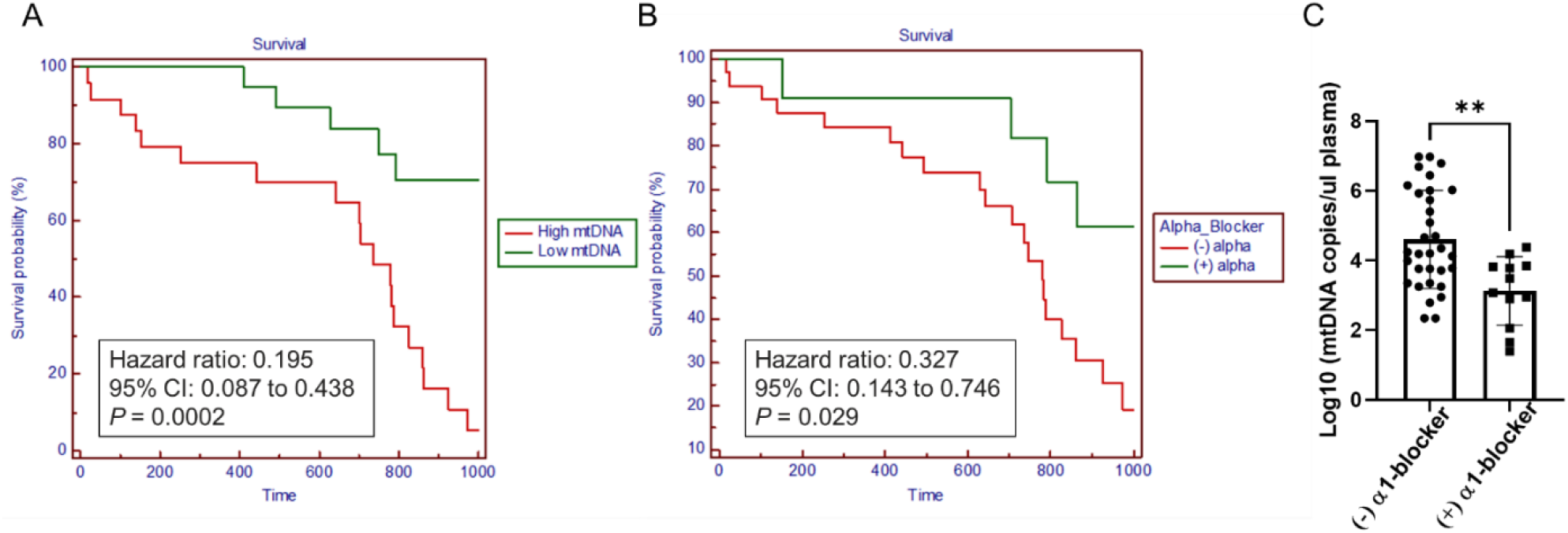
IPF patient taking α1 adrenoreceptor antagonists show reduced plasma mtDNA. (**A**) Kaplan-Meier plot for all-cause mortality reveals a significant survival benefit (a hazard ratio of 0.20, *P* = 0.0002) for subjects with a plasma MT-ATP6 concentration less than or equal to 3.785 log10 copies/µL (“Low mtDNA” group with green line, n = 20) relative to those with a plasma MT-ATP6 concentration greater than 3.785 log10 copies/µl (“High mtDNA” group with red line, n=24). (**B**) Kaplan-Meier analysis comparing patients prescribed an α1 adrenergic receptor antagonist (“(+) alpha” group with green line, n = 12) to those not on such therapy (“(-) alpha” group with red line, n = 32) showed a hazard ratio for all-cause mortality of 0.33 (*P* = 0.029). (**C**) Relative to plasma samples obtained from IPF patients who were not taking an α1 adrenoreceptor antagonist (alpha blocker) at enrollment (left), the mean concentration of MT-ATP6 was significantly decreased in plasma samples obtained from IPF subjects who were taking such drugs (right) (*P* = 0.0018). Data are presented graphically as log base 10 of the raw values of MT-ATP6 copies per microliter of plasma with mean (±SEM). CI: confidence interval. IPF: idiopathic pulmonary fibrosis. mtDNA: mitochondrial DNA. Time: days.

## DISCUSSION

This study lends new insight into the mechanisms through which α1 adrenoreceptor antagonism exerts its antifibrotic benefit in the lung. Specifically, using the bleomycin model of lung fibrosis we show that fibrosis-promoting NA is at least partially derived from local adrenergic nerves, that experimentally induced fibrosis involves the expansion of an αSMA+ population expressing ADRA1D, and that α1 adrenoreceptor antagonism with terazosin treatment reduces both the accumulation of αSMA+ cells in parenchymal regions as well as concentrations of extracellular mtDNA. In a parallel set of experiments, we show that NHLFs co-stimulated with TGFβ1-and NA express ADRA1D and respond to terazosin in a cell autonomous fashion by reducing both the expression of *ACTA2* and the accumulation of mtDNA. Finally, we show that the reduction in all-cause mortality that is seen in IPF patients taking α1 adrenoreceptor antagonists is paralleled by a reduction in circulating mtDNA. When viewed in combination, these findings suggest that α1 adrenoreceptor antagonism’s antifibrotic benefit involves a previously undescribed mechanism wherein nerve derived NA interacts with ADRA1D on lung fibroblasts to cause an activated state characterized by αSMA expression and mtDNA release. These findings carry great ramifications both for the relatively rare condition of IPF as well as for numerous conditions characterized by adrenergic overactivation and fibrotic remodeling.

The first novel finding of this study is the implication of NA derived from local, and not systemic, sources in the pathogenesis of pulmonary fibrosis. This conclusion is supported by the finding that locally delivered 6-OHDA results in a specific ablation of TH+ nerves and NA in the lung but not peripheral organs. It is also supported by the observation that ablation of a systemic source of NA, the adrenal glands, did not appreciably impact fibrosis in this model. While other sources of lung NA are possible, this possibility is unlikely. It is also possible that pulmonary accumulation of NA results from inadequate expression or function of catecholamine recycling machinery such as Solute Carrier Family 6 Member 2 (SLC6A2) and the flavoenzyme monoamine oxidase A (MAOA). These possibilities are not mutually exclusive and raise additional opportunities for investigation in this regard. As lung nerves do appear to be implicated, additional areas of study should focus on whether it is adrenergic nerves in the airway or blood vessels that exert this effect, and how non-adrenergic substances derived from sympathetic nerves such as Dopamine and Neuropeptide Y are involved. Finally, the contribution of nerve-derived NA to fibrotic lung disease raises the possibility of targeting local NA either through direct neuromodulation, inhalational administration of agents targeting the ADRA2A receptor to impede presynaptic release or postsynaptic receptor activation. Further work will be required to ascertain the benefit of these approaches.

The present study also raises the exciting possibility of repurposing orally available α1 adrenoreceptor antagonists that are FDA approved for other indications for the treatment of IPF. While studies have demonstrated its therapeutic potential of this drug class for the treatment of acute inflammatory lung diseases (43, 46), its benefit for chronic fibrotic conditions such as IPF has yet to be explored. In our work we show that αSMA+ fibroblasts express ADRA1D and are reduced by terazosin treatment in the bleomycin mouse model and in cultured human cells. This observation raises the intriguing possibility that the antifibrotic mechanism of α1 adrenoreceptors is mediated via direct inhibition of fibroblast activation, potentially through ADRA1D. Because Terazosin has been reported to be relatively specific for the ADRA1D receptor subtype (47) it is likely that its benefit is mediated through this receptor; however, further studies using cell specific knockout mice and competitive binding assays will be required to more fully understand this biology and leverage it for clinical benefit (48).

A particularly provocative and exciting aspect of our work is the finding that α1 adrenoreceptor antagonism mitigates the extracellular accumulation of mtDNA. This finding is in line with work in other realms where terazosin stimulates cellular resistance to stress via phosphoglycerate kinase 1 and heat shock protein 90 to improve cellular bioenergetics and mitigate mitochondrial DNA damage (37). In our study, α1 adrenoreceptor antagonism attenuated stress-induced extracellular mtDNA accumulation in rodent models, cultured NHLFs, and IPF patients suggesting a similar mechanism may be at play. In terms of the connection to fibrosis, because mtDNA stimulates a feedforward loop through which fibroblasts induce autoactivation by internalizing mtDNA to activate cytosolic DNA sensors such as TLR9 (21) and cGAS/STING (36), our finding suggests that α1 adrenoreceptor antagonism might interrupt this circuit. If true, this conjecture frames α1 adrenoreceptor antagonism as a new form of neuro-innate signaling and suggests that circulating mtDNA concentrations could be used to monitor drug efficacy and treatment response. Additional work using transgenic mouse modeling and dedicated in vitro studies will be required to more fully explore this connect, as will larger prospective studies of patients with IPF. Nevertheless, these findings carry significance for the relatively rare condition of IPF as well as for numerous cross organ conditions of adrenergic overactivation, inflammatory remodeling and mtDNA such as non-alcoholic hepatic steatosis (33, 49), cardiac fibrosis (50), obesity (51, 52), ARDS (43, 53), and COVID-19 (46, 54), and suggest a convergent mechanism for divergent disease states.

While important, our study leaves unanswered several questions that constitute the basis for additional study. First, we have not explored whether cell populations beyond fibroblasts are effectors in the pathogenesis of NA-mediated lung fibrosis. Given the widespread expression of adrenoreceptors on resident and recruited cells it is likely that other receptors and processes are involved. Second, while our data suggest ADRA1D as the mediator of NA’s stimulatory effects on fibroblasts, we have not performed the overexpression and knockdown studies required to support this claim. Furthermore, while the available data suggest that α1 adrenoreceptor antagonism exerts cell autonomous effects on fibroblast activation and mtDNA release, we have not ruled out paracrine processes and additional sources of mtDNA. Indeed, given the pleiotropic processes involved in fibrosis it is possible and in fact likely that additional mechanisms are involved. Finally, the mechanism through which α1 adrenoreceptor antagonism reduces mtDNA in IPF patients will require prospective studies.

In conclusion, our work links nerve-derived NA and α1 adrenoreceptor antagonism with mtDNA accumulation and lung fibrogenesis in mouse models, cultured cells, and humans with IPF. Further study of this neuro-innate connection may yield new avenues for investigation in the clinical and basic science realms.

## Data Availability

All data produced in the present study are available upon reasonable request to the authors.

## AUTHOR CONTRIBUTIONS

GI conducted the investigation, methodology, formal analysis, visualization, wrote and edited the manuscript. XP, JM, SW, CP, and AL performed the investigation. GH, AG, SY, and HS conducted the methodology and conceptualization. CR and ELH provided conceptualization, resources, visualization, supervision, project administration, and funding acquisition and wrote and edited the manuscript.

## ACKNOWLEDGEMENTS

We thank Guanling Huang at Cedar Sinai, CA, for intellectual support of the project. We kindly thank all of our IPF patients who generously donated their time and biospecimens to TYLR.

