## Supplementary figures for "α1 adrenoreceptor antagonism mitigates extracellular mitochondrial DNA accumulation in lung fibrosis models and in patients with IPF"

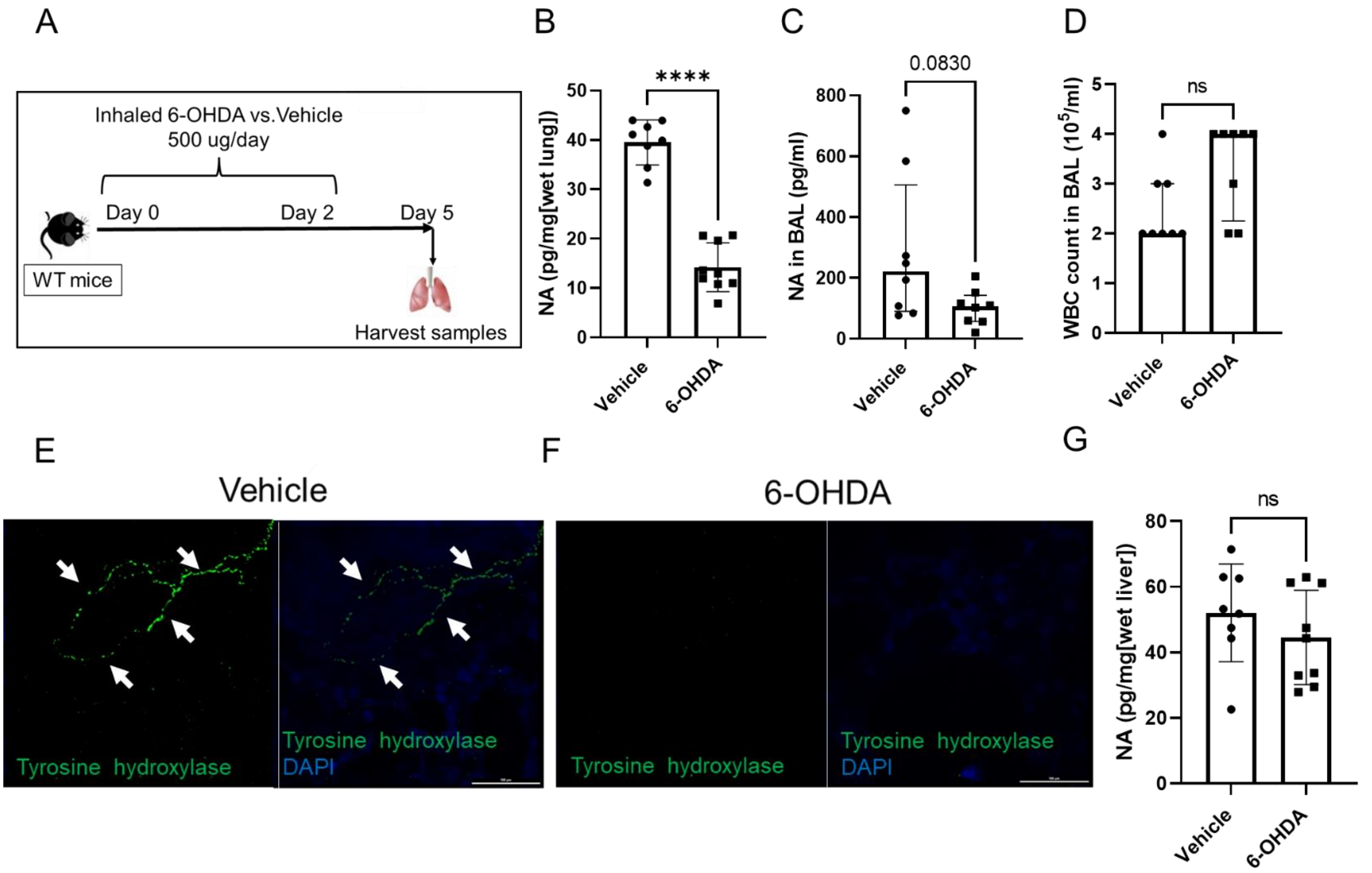


**Supplementary figure 1**: **Inhaled 6-OHDA depletes local adrenergic nerves and decreases noradrenaline in the lung.** (**A**) WT mice were treated with either L-ascorbic acid (vehicle) or inhaled 6-OHDA from Day 0 through 2 and sacrificed at the 5-day time point. (**B** and **C**) Comparison of NA concentrations in mice treated with vehicle (left) and 6-OHDA (right) revealed that relative to vehicle-challenged groups, the NA concentration was substantially decreased in lung of 6-OHDA treated mice (**B**, $P < 0.0001$). Concentrations of BAL NA (pg/ml) approached but did not reach significance in 6-OHDA treated animals (**C**, $P = 0.0830$). (**D**) Mice challenged with inhaled 6-OHDA (right) showed unchanged WBC count in BAL relative to mice challenged with vehicle (left) ($P = 0.1026$). (**E** and **F**) Immunofluorescence detection of TH (green) and DAPI (blue) in lungs treated with vehicle (**E**) and inhaled 6-OHDA (**F**). In the vehicle image in **E**, white arrows indicate TH+ linear structures were localized around airway. In the 6-OHDA image in **F**, TH+ linear structures were undetectable. (**G**) Comparison of liver NA concentrations in WT mice challenged with vehicle (left) and inhaled 6-OHDA (right) revealed that relative to vehicle-challenged groups, the hepatic NA content was unchanged ($P = 0.3092$) at the 5-day time point in mice challenged with inhaled 6-OHDA. ****$P < 0.0001$. Data are shown as mean ± SEM or median ± IQR. BAL: bronchoalveolar lavage. NA: noradrenaline. 6-OHDA: 6-hydroxydopamine. TH: tyrosine hydroxylase. WBC: white blood cell. WT: wild type. Scale bar = 100 microns.

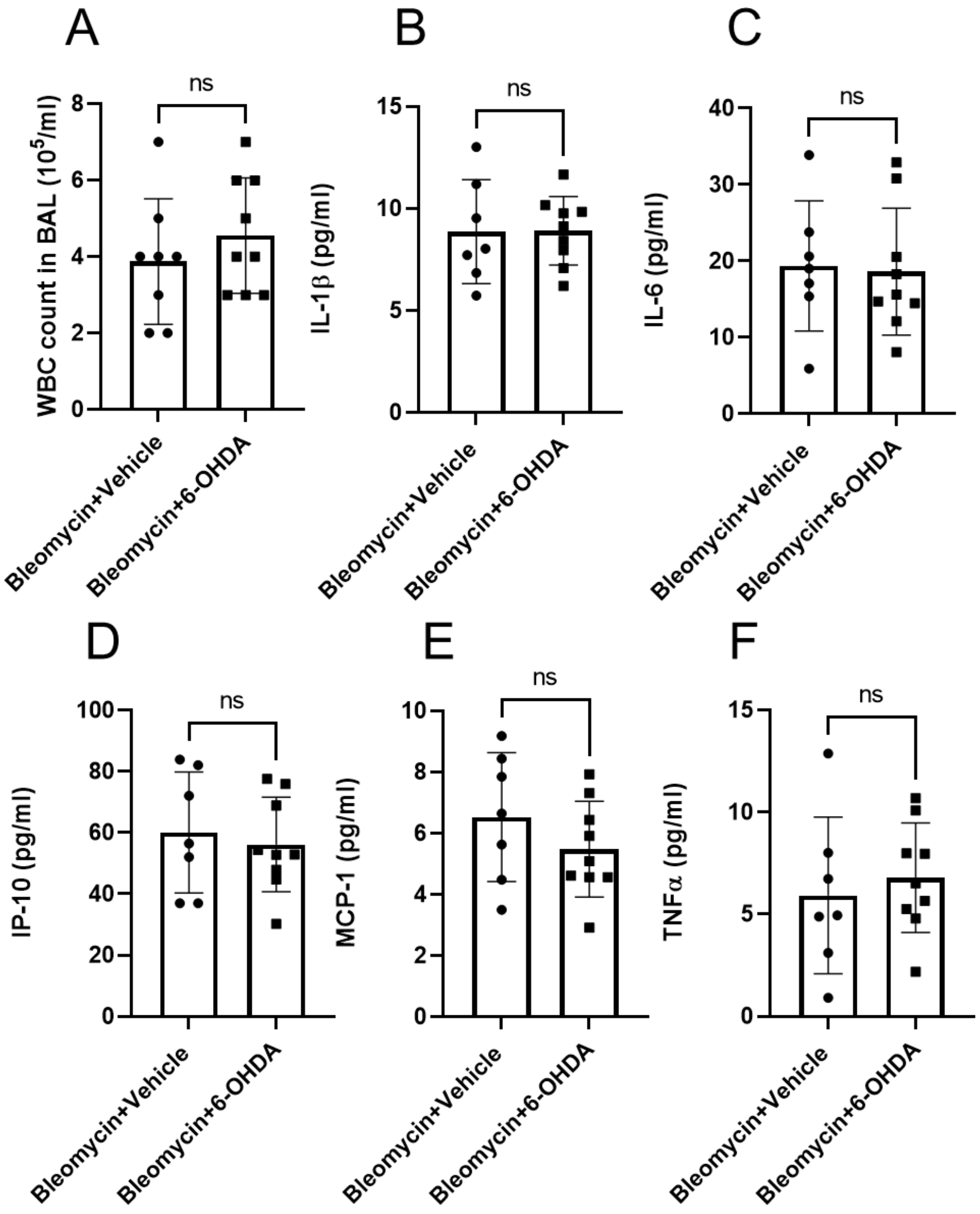


**Supplementary figure 2: Comparison of BAL immune parameters following inhalation of 6-OHDA.** (**A**) Following bleomycin challenge, mice treated with inhaled 6-OHDA (right) showed unchanged BAL WBC count relative to mice treated with vehicle (left) ($P$ = 0.3872). (**B**-**F**) There was no difference in BAL concentrations of IL-1β (**B**, $P$ = 0.9619), IL-6 (**C**, $P$ = 0.8607), IP-10 (**D**, $P$ = 0.6603), MCP-1 (**E**, $P$ = 0.2703), and TNFα (**F**, $P$ = 0.6031) between bleomycin challenged mice with inhaled vehicle control and with 6-OHDA. 6-OHDA: 6-hydroxydopamine. BAL: bronchoalveolar lavage. WBC: white blood cell. Data shown as mean ± SEM.

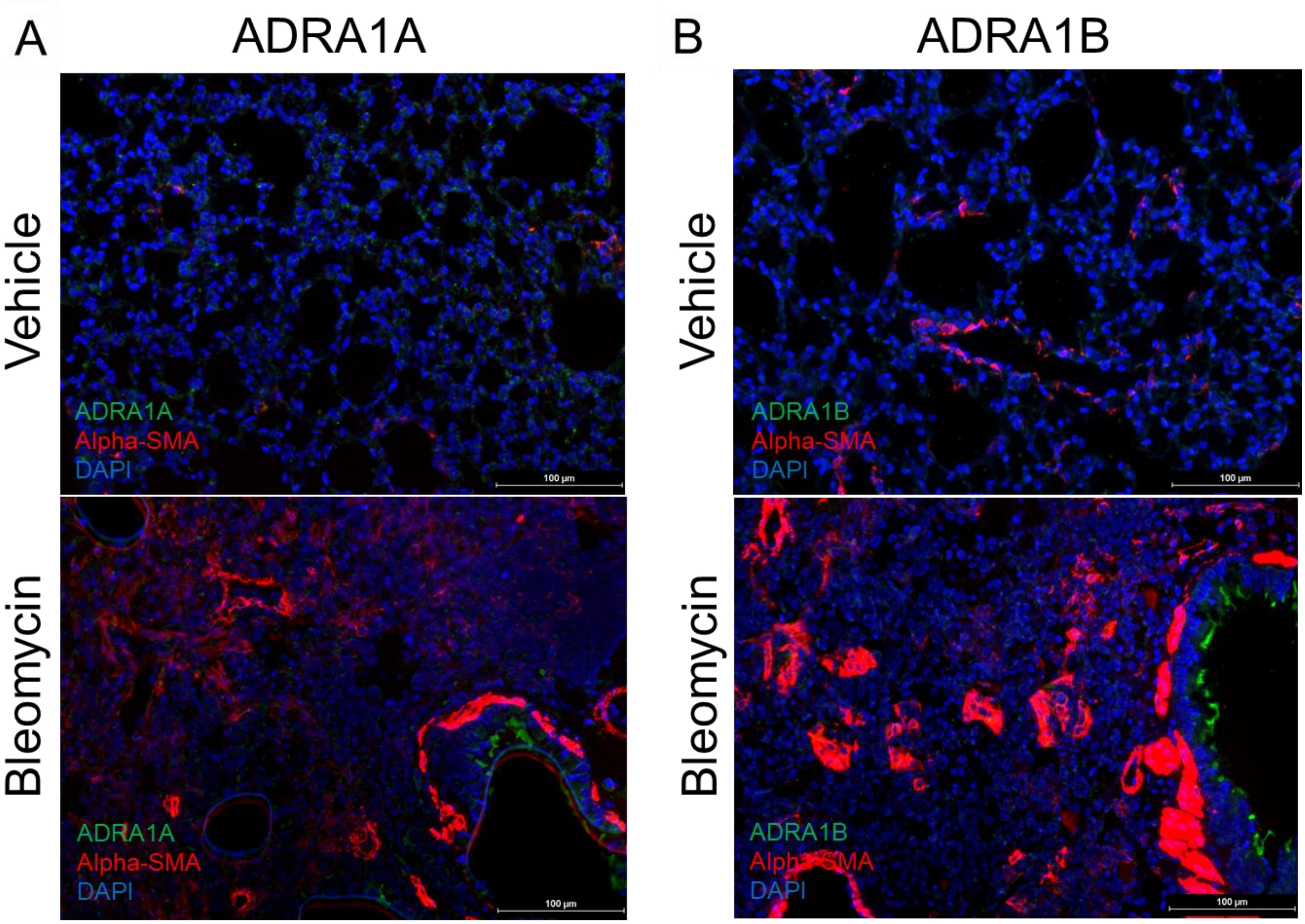


**Supplementary figure 3: Geospatial localization of cells expressing ADRA1A, ADRA1B, and αSMA in bleomycin or vehicle challenged lungs:** (**A** and **B**) Immunofluorescence detection of (**A**) ADRA1A or (**B**) ADRA1B (green), αSMA (red), and DAPI (blue) in both control and bleomycin challenged lungs revealed the absence of ADRA1A (**A**) or ADRA1B (**B**) expressing αSMA positive cells. αSMA: α-smooth muscle actin. Scale bar = 100 microns.

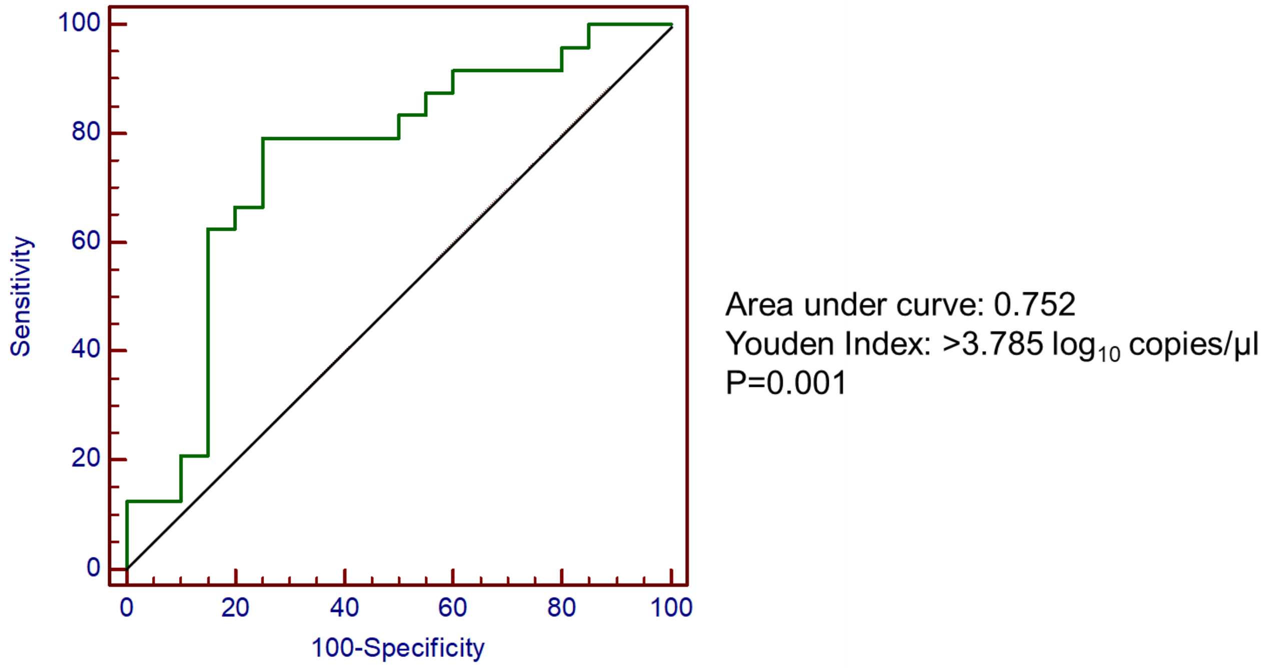


**Supplementary figure 4:** Receiver operating characteristic curve analysis of the Yale-ILD cohort was used to determine the mtDNA cutoff. mtDNA: mitochondrial DNA.
